# “It’s no use saying it in English”: A qualitative study exploring community leaders’ perceptions of the challenges and opportunities with translating and interpreting COVID-19 related public health messaging to reach ethnic minorities in Australia

**DOI:** 10.1101/2023.03.23.23287618

**Authors:** Holly Seale, Ben Harris-Roxas, Anita Heywood, Ikram Abdi, Abela Mahimbo, Lisa Woodland, Emily Waller

## Abstract

**Background:** The Australian Government implemented a range of public health response strategies and communication approaches to reduce the spread of COVID-19; however, concerns have been raised around a failure to consider culturally and linguistically diverse (CaLD) communities sufficiently in these processes. This research aimed to understand the factors that have impacted COVID-19 communication and engagement efforts during the pandemic from the perspective of key CaLD community stakeholders and community members. A further aim was to understand the processes that could be adopted to support future communication strategies, including the promotion of pandemic-related vaccines.

**Approach:** This study included 29 key informant interviews with community and faith-based leaders in New South Wales, Australia.

**Results:** The overwhelming message from community leaders was a sense of shared responsibility between their organisations and the government in communicating pertinent and accurate COVID-19 related information to CaLD communities. They expressed a sense of duty to keep their community members safe. While acknowledging this shared responsibility, community leaders and others shouldered significant costs related to resources and time that need to be acknowledged by governments in preparing for future disease outbreaks. They felt that governments should consider: 1) improving communication between governments and CaLD organisations; 2) responding to the specific CaLD needs with greater agility; 3) foregrounding social media in their communication strategy; 4) reinvesting in local public health units to know their population; 5) investing in a health ambassadors model program; 6) preparing a hybrid model of translators/interpreters to fill the gap; and, 7) reimagining vaccine information campaigns to better target CaLD communities.

**Conclusion:** Given the technical details about the COVID-19 virus conveyed in government information campaigns and the media, ensuring the most vulnerable populations, including people from CaLD backgrounds, access clear, concise and timely public health messaging from both governments and community organisations requires further attention.

## INTRODUCTION

Much has been written about the impact that the global COVID-19 pandemic has had on vulnerable communities, particularly ethnic minority populations (the convention in Australia is to refer to such groups as culturally and linguistically diverse populations (CaLD), and thus we will be using this term throughout the paper) [1-5]. Numerous studies have documented the heightened risk from SARS-CoV-2 and the disparity in COVID-19 cases and deaths amongst a diverse group of migrants, refugees, and asylum seekers and other ethnic groups [6,7]. Evidence suggests that in some settings, these at-risk groups of migrants have been excluded from the national response [2,5]. Several factors have contributed to this, including a lack of consideration and tailoring given to the needs of CaLD population groups within pandemic response strategies and inaction amongst governments to ensure that resources are not only available in wide range of languages but are also culturally relevant [6]. Reviews focused on the quality of COVID-19 communication materials have highlighted either a lack of resources on disease prevention for some migrant population groups (people in refugee camps or informal settlements), limitations in the number of languages information was translated into, or issues with the complexity (readability, understandability, actionability) of information or issues with the quality of the translations [2, 8-9]. Studies from Denmark, Australia, Canada, and the US have documented delays in the availability of official guidance in relevant languages for the respective countries and issues with the approaches used to disseminate resources into CaLD communities, with a particular focus on groups with lower language and literacy levels, or communities with oral languages [3, 10-11]. These issues are heightened amongst older people from CaLD backgrounds, which can be attributed to their inability to search for health information online and a communication gap between the adults from CaLD backgrounds and the public health information dissemination system [12]. CaLD populations can be left behind in their access to and understanding of recommendations, compounded by cultural discordance and mistrust of health institutions [13].

In Australia, English is the dominant language; however, many people speak a language other than English within their families and communities. There are over 200 languages spoken in Australia, including over 50 actively spoken Australian Indigenous languages. The availability of information in other languages often depends on the size of the community, the level of community infrastructure, and engagement of the community with health services around other health-related issues. In Australia, speakers of languages with sizeable numbers of practitioners – Mandarin, Arabic, Vietnamese, Cantonese, or Greek, may glean information from national- or community-based radio stations and newspapers, among other modes of communication. But other communities are classified as linguistic minorities which do not have this level of community infrastructure. Research has found that people from CaLD communities identify strategic stakeholders (referred to in this paper as ‘information intermediaries’) for disseminating information within groups [14,15]. Information intermediaries could include staff from community organisations, community or religious leaders, bilingual case or youth workers or ‘natural’ leaders (for example, a person who has completed medical training but does not practice in Australia) across the age spectrum. Through influencing the flow of information, information intermediaries shape and inform their community’s reality and knowledge in a culturally appropriate and salient manner.

While guidelines and reports highlight the crucial role of these ‘information intermediaries’ in supporting communities during the COVID-19 pandemic, less is known about the experiences of these information intermediaries in terms of their perceptions of the COVID-19 response, the needs of their communities and their experiences in ‘bridging the gap’ between levels of government and the community in communicating pertinent COVID-19 information. Given the concerns about the Australian Government insufficiently considering the needs of CaLD communities in their public health response strategies and communication approaches to reduce the spread of COVID-19, this research aimed to understand the factors that have impacted COVID-19 communication and engagement efforts during the pandemic from the perspective of key CaLD community stakeholders and community members. A further aim was to understand the processes that could be adopted to support future communication strategies, including the promotion of pandemic-related vaccines, as proposed by CaLD information intermediaries.

## METHODS

Semi-structured, in-depth phone interviews with people who self-identified as community or religious leaders, which were approximately 30-40 minutes, were undertaken between March and September 2021. The focus was on CaLD communities that include people born in mainly non-English-speaking countries, and where English is not the primary language spoken at home [14]. The Human Research Ethics Advisory Panel at the University of New South Wales reviewed and approved this study (HC200776). Informed verbal consent was collected from all participants.

### Sampling

We worked with a range of key organisations including migrant resource centres, refugee health services, settlement services, peak bodies and community-based organisations, and government organisations, to promote the study opportunity to known leaders within their networks. Information was sent out via these networks, with potential participants encouraged to connect with the research team for further information or to express interest. Participants were only included in the study when full verbal consent had been received. This study did not collect any identifiable personal information from the participants. Recruitment for this study was targeted to those living within New South Wales, Australia.

### Data collection and analysis

An interview guide was developed and reviewed by the researchers to identify critical areas of interest for the study. The questions related to the following topics: perspectives towards the current communication approach being used by the government, factors affecting communication and engagement with CaLD communities, the communication roles and influences of different agencies, and suggested options that could be adopted to enhance communication and engagement around the COVID-19 vaccine program. Questions were asked in an open-ended manner to allow room for expansion [16].

Thematic analysis is a method for identifying themes and patterns of meaning across a dataset in relation to a research question; possibility the most widely used qualitative method of data analysis [17]. A six-step thematic analysis qualitative framework developed by Braun and Clarke guided data analysis [18]. The first investigator reviewed a small selection of transcripts for early themes and concepts, from which the preliminary coding scheme was constructed, and then a second investigator coded all transcripts, revising the scheme iteratively to reflect emergent themes from interview responses. QSR International’s NVivo 12 qualitative data analysis software was used to code all transcripts, categorise the data and facilitate comparison of participant views. A third investigator independently coded at least 20% randomly selected interviews to ensure coding consistency. The investigators convened to share their categories; any discrepancies found were resolved through discussion and negotiated consensus. Data analyses and interpretation were iterative, and all investigators participated in this process, to identify and agree upon emergent themes, and discuss their face validity. Two of the seven team members identify as coming from CaLD communities, and their input was invaluable in ensuring relevancy to broader CaLD communities.

## RESULTS

Twenty-nine interviews were undertaken with community and faith leaders based in New South Wales. The characteristics of the participants and the interviews are described in Appendix 1 using the CORE-Q reporting format [19]. The themes identified that apply to how government communication with multicultural communities include: 1) *“They know nothing”:* understanding the language and generational divide affecting CaLD populations; 2) *“I expected a lot more engagement”:* critically assessing the government’s engagement with CaLD communities; 3) *“How much perfection do we need?”*: expressing support for the government’s engagement with the CaLD community; 4) *“Bridging the gap”*: CaLD communities leaders stepping up; 5) *“With some translation I would cringe”:* quality and scope of translations matter; 6) *“Most of them are tuning into overseas stations”:* relying on overseas health messaging among CaLD communities in Australia; 7) *“You can’t reach them by the traditional way”:* harnessing social media; and 8) “*The people really want to know the truth”:* challenges of communicating complex medical information to CaLD communities. These themes are described in detail below.

### 1. “They know nothing”: Understanding the language and generational divide that impacts CaLD populations

While the CaLD communities are important and valued members in Australian society, their diversity and heterogeneity present unique challenges in ensuring accurate public health messaging is accessible for all. One of the community leaders simply stated: “Within the non-English speaking communities, there are some people that require more support than others” [Community Leader (CL) 06]. Two of the most notable challenges highlighted by the community leaders who participated in this research was: 1) an English language divide, where basic English proficiency was not sufficient – or absent - to fully understand the rapidly evolving public health messaging; and 2) a generational divide, where accessibility to quality translations of public health information was limited, especially in the absence of internet use and social media proficiency.

Our participants noted that some CaLD community members have basic English language proficiency, though many depend on community language newspapers or community language radio stations (such as SBS [Special Broadcasting Service] in Australia), rather than mainstream English-language news outlets [CL 03, 16, 21]. “The majority of migrants [only] have basic English language skills, so they need a lot of support. They need the information to be translated…so they can read it and understand it well” [CL 21]. Another community leader noted, “They stick to their own people…They’re quite happy with their own language among their own people. It’s easier for them to receive the information in [their community language] rather than in English” [CL 16]. Furthermore, it was reported that some people from CaLD backgrounds do not have high levels of education, and may even be illiterate in their own language, and require complex public health information to be distilled into more simple and clear instructions [CL 04, 05, 15].

Ageing populations with the CaLD communities was also raised several times by community leaders as a major factor in considering how to disseminate public health information. Besides language, they are impacted by a digital divide – unable to navigate the internet to access critical public health information, despite some community training efforts [CL 03]. While some elderly people from CaLD backgrounds have family in Australia able to offer support, others are isolated and have limited outreach avenues [CL 17]. “How much do [older people from CaLD backgrounds] know about what’s going on?…[W]hen I would go for walks in my [neighborhood], I’ve been curious to ask [older] people, what do they know? They know nothing” [CL 17].

Older people from CaLD backgrounds need special consideration, especially in historically insular communities where CaLD communities migrated to areas for manufacturing industry jobs nearly half a century ago. As a result, there emerged a large migrant community that meant immersing themselves in the English language was unnecessary. “When they were coming 40 - 50 years ago, there was no need for them to know any English at all, they would get a job and work for decades, without them learning too much English. Then because there was so many of them, they didn’t need to learn English” [CL 11]. However, as the population has aged, accessing information has become more challenging, relying on overseas information in their community language, which may not bear much use in the Australian context [CL 11]. The intersection of how people from CALD backgrounds, English translations and interpretation, generational differences, the role of the internet, and technical public health information played out during the pandemic in Australia will be explored in more detail in the themes that follow.

### 2. “I expected a lot more engagement”: Critically assessing the government’s outreach to CaLD communities

Feedback from community leaders was largely critical of the government’s COVID-19 engagement and communication efforts with CaLD communities. There was a general sense of frustration with how essential public health information was disseminated to CaLD communities, with many community leaders pointing to limited resources that were translated into languages other than English, poor quality translations (see theme below on “Quality of translations matter”), and lack of outreach to the community organisations. One respondent described the dissemination of government public health messaging as “poorly” [CL 11]. Another leader noted, “There were no information reaching the multicultural community, especially, the non-English speaking backgrounds” [CL 06]. Another observed that information about vaccines was limited. “I did not see much engagement with the community about the vaccination, I must say…I [would have] expected a lot more engagement; a lot more” [CL 17]. This respondent pointed to the daily eleven o’clock TV COVID-19 news updates as the most notable engagement with community [CL 17]; though such outreach would only reach a section of the community with higher levels of English proficiency.

Participants did acknowledge that while the beginning of the pandemic was worse in terms of accessing information, communication did improve as the pandemic unfurled and new learning emerged. “[Information] wasn’t that great at the beginning. There was a lot of individual work of the community to convey the message. The government wasn’t doing much in that regard” [CL 21]. Though participants acknowledged that during the second lockdown, availability of information in community languages improved. However, there remained an information gap that was filled by community members stepping in to translate key messages, such as general public health statistics, lockdown rules, and other relevant updates. “I knew a few people who are doing it themselves, without pay, without anything, [with] nothing, just the individual - They were volunteering to do it” [CL 21]. While communication with CaLD communities were enhanced as the pandemic progressed, several participants cited a lack of a coherent strategy targeting the diverse CaLD populations and an over-reliance on community organisations to lead in the delivery of public health messages in different languages continued throughout the pandemic [CL 20].

### 3. “How much perfection do we need?”: Expressing support for the government’s engagement with the CaLD community

Criticism or frustration at the government’s outreach efforts was not shared universally across our interviewees. Some community leaders expressed satisfaction with how messages about COVID-19 and translated materials were distributed to CALD communities. “We are fairly satisfied. Whatever messages we get, we share with our community through our [online newspaper]…[W]e [also] motivate the people in our community to organise [through] Zoom meeting[s]” [CL 02]. Online forum meetings was highlighted as an important mode for dispersed community members across the country to come together and share information regarding COVID-19. Similarly, others less critical of the government’s response, suggested there needs to be limits on the number of different languages into which the government is expected to translate information. “I just don’t understand how many languages we need to translate and how much perfection do we need? It’s an emergency situation” [CL 01]. This respondent also noted that compared to other places in the world, the ability for the government to cater to a wide-range of languages should be commended: “Somebody came up with so many translated documents…It was the best thing that has happened” [CL 01].

Other respondents seemed to suggest that ensuring translated documents are created and distributed must be a shared responsibility between the government and the community, and that the community has a role to play in facilitating the knowledge transfer. Such cooperation may include volunteering to translate documents on behalf of the government. “Many times we have taken those documents and got a person to translate it…and then distribute it. What’s the big deal about it? I can…sit down for half an hour and [translate] it” [CL 01]. However, while some respondents expressed ambivalence about the task to translate certain documents, others took issue with the amount of time and skill required to do this work without funding or support.

### 4. “Bridging the gap”: CaLD communities leaders and organisations stepping up

Several community leaders used the phrase ‘bridging the gap’ or ‘filling the gap’ when describing their organisations’ role in disseminating public health messaging to their relevant communities [CL 09, 20, 21]. Several participants noted that the act of translating, as well as the act of determining in which networks to circulate the information, was a time-consuming endeavour, especially those organisations where funding was not available for formal translations [CL 03]. Leaders representing communities from South Sudan, China, Indonesia, and Iraq, among others, shouldered extra tasks to ensure that the government’s COVID-19 information reached their respective communities. “There was a lot of individual work of the community to convey the message. The government wasn’t doing much in that regard. Only now with the second lockdown, we’re starting to see more information in community languages” [CL 21]. Some of these community leaders did the translating and interpreting themselves, using their bilingual fluency (some were fluent in up to five languages) to guarantee messaging reached the most vulnerable populations, while others engaged colleagues and international students to assist in translations and interpretations [CL 06, 09].

At the start of the pandemic, some community leaders conducted outreach to their communities by creating social media pages, video and radio content in their community language and sharing them online [CL 16]. The messages in the videos contained basic public health information for avoiding the virus, ranging from how to properly wash hands to social distancing etiquette.

While the radio segments were broadcasted to the wider population, the videos were distributed through a network of smaller organised groups, such a women’s groups, etc. [CL 16]. Another leader created a CaLD-community specific COVID Emergency Facebook page where awareness-raising and information sharing was accessible amongst community members [CL 06].

While several community leaders stepped up to ensure public health communication had a further reach, many also acknowledged that this engagement came with a significant time and resource cost to the individual and/or the organisation [CL 21, see the second theme above]. One leader noted that they received several phone calls a day from community members requesting clarification on a number of COVID-related issues, such as vaccines and lockdown rules. “Our community start[ed] to ask more questions…I receive[d] personally more than - I’m not exaggerating - 50 to 80 calls every day, people asking me to clarify a few things, which I’m ready to do it. I’m happy to [give people] the facts” [CL 20]. Fielding phone calls, translating documents, sending messages, and uploading original videos were among the many modes of communication community leaders initiated to share pandemic-related public health information.

The suite of communication strategies employed by the community leaders were not without their challenges. For example, when preparing for a radio segment, air time is extremely limited, necessitating that important, and sometimes complex, public health content had to be distilled to a concise five-minute timeframe [CL 16]. Another leader noted that trawling through all of the public health data and information to determine what the most pressing messages required translation and which dissemination channels to use was time-consuming work [CL 03]. Similarly, reviewing the onslaught of COVID-related messages on the WhatsApp platform was tedious, and community organisation needed support to navigate the information. “I feel that if we can have people who actually looked through [the material], and see what’s relevant and convey that information to our communities, I think that would work” [CL 03]. Such efforts took preparation and translation time of the respective community leaders, which was not necessarily reimbursed or part of their roles, though most all community leaders we spoke to indicated they were prepared to shoulder such work given the extraordinary circumstances of the pandemic and the desire to protect their communities. However, greater communication between governments and CaLD community organisations to discuss this shared responsibility and how it can be properly resourced for future pandemics, and public health campaigns more broadly, is imperative.

### 5. “With some translation I would cringe”: Quality and scope of translations matter

Community leaders emphasised the importance of having high quality translated material in order to reach the large swaths of their community where English is neither spoken nor proficient. “It’s no use saying it in English” was the response of one community leader when highlighting the critical need for translated material [CL 15]. Furthermore, participants raised several important points related to the scope of the translations, in terms of the languages in which public health information is translated, but also the nuances *within* languages and how critical it is to take into consideration the wide range of dialects. Finally, several community leaders highlighted challenges and opportunities related to the process of translating public health messages, whether through accredited bilingual community educators or through more informal mechanisms lead by community organisations.

#### The scope of translations

One of the most important decisions governments make in relation to communicating to CaLD communities is the languages in which outreach material are translated. Whilst governments may prioritise translations based on census data indicating the highest proportions of languages spoken in the home, community leaders noted that some groups felt ostracised when their language is not part of the official list of translated language. “There’s definitely been other groups that have felt that they’ve slipped in between the cracks, and it was that whole premise of, ‘Do you translate resources into the languages where there is the vast amount of people? Or do you go for those languages where maybe there are groups who are at heightened risk or have particular needs?’ [CL 09]. Partcipants signaled that smaller CaLD communities may be at greater risk of pandemic-related harms because of the lack of community infrastructure around them.

As noted in the first theme above, translated material needs to take into account the educational background of their target audiences. “You’re not translating to people who are educated and work…You have to be very clear [when you translate]” [CL 04, echoed by CL 15]. In some cases, translators and interpreters had to deviate from exact translations to make new concepts understandable, such as the use of QR codes to check-in to venues. Others noted that the technical language associated with some public health information may be challenging for the average person, and especially those who were not proficient in English. “If I have difficulties understanding what they’re talking about because they’re using the language that is very technical, the jargon and all that. What about the common person?…Whether it’s COVID or non-COVID, this is the problem” [CL 15].

Another important translation issue that was consistently raised across our community leaders was the need for greater attention to the dialects *within* languages. Translations tend to be in the dominant languages of established CaLD communities. However, several community leaders advocated for additional resources to reach more diverse language groups, particularly in Indian and Middle-eastern communities [CL 03, 04 10]. As one community leader noted, even within seemingly cohesive CaLD communities, there is internal diversity amongst the dialects [CL 04]. A community leader representing the Indian community in Sydney noted: “We’re such a large community and we speak so many different languages…that is the point I would like to stress” [CL 03]. This participant reported that most translated resources are in Hindi, Tamil or Punjabi. However, they continued: “[t]here are seven other languages that are spoken [in the Indian diaspora in Australia] which are not addressed…Although it is widely spoken here, not everybody understands Hindi. That’s where we step in, because we can speak other languages. We use our ambassadors to speak for example, in Gujarati, in Bengali…We use them as ambassadors to communicate the information in their native language” [CL 03]. This particular community organisation filled a gap where additional translation and interpretations services were needed to communicate to community members speaking minority languages. This sentiment was echoed by an Arabic-speaking community leader, who noted the critical importance of sharing information in several different dialects, whether Chaldean, Ashuri, or Syrian, among others [CL 10]. Another community leader re-enforced the importance of knowing which dialect will suit the audience. “For example, [i]n the Arab world, we have 18 countries…If we’re reading a book, it’s all the same language…, but the [spoken] dialect is very different…When I speak to Iraqis with a group of Sudanese, Lebanese, Tunisians, Kuwaitis or people from Dubai, I have to modify my language. It’s not the same dialect…[D]ue to the needs within the Arabic community, you’ll need to adjust your language” [CL 04]. This observation was reflected by another community member: “The problem is you cannot just translate an English text to an Arabic text. It doesn’t reflect the exact meaning or the essence of the message’ [CL 19]. While another community member echoed this suggestion, they noted as well that only educated members of the Arabic community will be able to read documents translated into Arabic [CL 15]. “I’m finding that if we only rely on the translation of documents, [it’s] a problem because it’s very hard to translate documents into plain Arabic, especially when we don’t even have it in plain English. In the Arabic language, if anyone from any of the Arab countries is to read it, that person has to be educated” [CL 15].

Challenges with accuracies *within* languages played out in Chinese communities as well. One community leader noted that most government communication on COVID-related health information was translated into simplified Chinese, the written language of mainland China that emerged in the mid-twentieth century to simplify the more complicated traditional Chinese characters as a way to promote literacy nationally [CL 09]. However, people from Hong Kong, Macau and Taiwan, for example, do not read simplified Chinese, they continue to read traditional Chinese. Furthermore, some migrants from Hong Kong may not be proficient in English despite the historical British colonial rule. CL 09 shared an example from a few years past when a local government council wanted information translated into simplified Chinese. When this community leader pushed back that some migrants only read traditional Chinese, the council responded: “‘Because the statistics show that the majority of the Chinese are from Mainland China, they go for simplified Chinese.’ Of course, as a member of a small-sized community, I can’t say too much…[but] I wanted to speak out on behalf of these people” [CL 09]. While this community leader later acknowledged improvements were made during the course of the pandemic, they did caution that checking translations for accuracy and the appropriateness of the translated language should remain a priority.

#### The process of translating public health material

Early in the pandemic, community leader noted that the process to translate COVID-19 health advice was too slow. “There was no information reaching the multicultural community, especially, the non-English speaking backgrounds” [CL 06]. Translation procedures could not keep up with the fast-paced shifts in messaging as the pandemic evolved, sometimes leaving days between a change in health advice and an official translation being circulated, at which point some advice was already outdated [CL 22]. “I think [it] took too long to communicate with the multicultural communities and to get our leaders involved…and [now] we have to be very quick and fast” [CL 15]. Efforts were undertaken to accelerate the translation and outreach processes as the pandemic progressed, both by government, especially through the use of bilingual community educators, and by community organisations (see the fourth theme above on ‘bridging the gap’).

The bilingual community educators, whom Local Health Districts within New South Wales (NSW) Health regularly engages were seen as an asset in bridging the government’s health advice with the community, providing an important service for communicating health advice to the wider community [CL 03]. “[Bilingual community educators are] great because they can speak the languages and they can communicate the information…They’re trained, so [their services] are very reliable and accurate. I think it’s good to employ bilingual community educators because the language proficiency may not be as good across the community” [CL 03]. This community leader noted that bilingual educators – as well as GPs – are often tapped to translate written resources for community organisations or speak to the media, and some feel these professionals should be utilised more for government translation and interpretation [CL 03]. “Why not use GPs to communicate this information? We use them. We also quite often are approached by the media, for example, radio, wanting to interview a GP. We do connect them with GPs who speak other languages” [CL 03]. The expanded role of informal information intermediaries is highlighted as a potential important component in future pandemic preparedness with CaLD communities.

One community leader emphasised the need for government to reinvest in community engagement, in knowing one’s population, rather than rely on formal modes of communication [CL 03]. There may be a missed opportunity to work with the people who are on the ground to disseminate information, rather than being over-reliant on the hard copy translations. Print material was especially flagged as being less useful. This participant bluntly noted: “People don’t read” [CL 03]. While this is certainly an overstatement, it does raise the possibility that other modes of communication should be foregrounded – social media in particular – for reaching a wider swath of the CaLD communities (see theme below on harnessing social media). And it further emphasises the need to focus on short, sharp bits of information, rather than expect community members to read through detailed health advice. The participant noted that the over-circulating of print material, especially through email, had the opposite effect of its intent. The “bombardment” of health information emails led to people ignoring them, rather than engaging with them [CL 03].

Another community leader noted the value of bilingual community educators reaches beyond the simple act of translating. Their intimate knowledge of the community and culture allows for a more nuanced translation that may take into account people’s level of understanding of Australian culture and English language ability, education level and/or years in which they have lived in Australia [CL 16]. There is also an undercurrent of trust between the bilingual community educators and the community that does not animate well from a generic translated document. One participant noted that while people from CaLD communities may have a basic understand of English, they “absorb” more in their community language[CL 16].

Where bilingual community educators could not fill the knowledge gap with CaLD communities, community organisations needed to step in. Community leaders discussed the various ways in which the translation process unfolds within their organisations. In some cases, it is simply the leader themselves who takes the initiative to translate and disseminate COVID-19 information. In other scenarios, there is a more thorough system of checks and balances, where the translator, editor and proof reader are all separate individuals [CL 04]. “Most of the material we translated [was] perfect because it will go through different channels. The person translating is not the same person proofreading it” [CL 04]. For some CaLD organisations, there is a further layer of check related to cultural or religious sensitivity [CL 15]. While less relevant for COVID-19, it is important for other health concerns, such as sexually transmitted infections [CL 04].

Community leaders expressed frustration at the quality of translations they were seeing in the community. “In shopping centers, for example, I would pass by a board with some translation and I would cringe sometimes because of the language. I’m not sure if they use Google translate…” [CL 04]. Another community leader commented similarly: “I did see that there has been some improvement [in the translated materials] over the year but I just hope that they will also have somebody to [double] check to see that it is actually the way it should be translated” [CL 09]. While these two community leaders did not specify if the translated materials were government endorsed translations, it does point to a general unease with the accuracy of public health information translations more generally. While there has been criticism about the quality of translated material, anecdotally, people have also pointed to the original messages in English being less clear. It’s not necessarily just the translations but that the original text wasn’t written in a way to make it easy to translate. However, in raising these concerns, another tension arose among our participants: whether translator and interpreters should act only if certified by the National Accreditation Authority for Translators and Interpreters (NAATI)? Assessment fees for individuals and institutions can range from hundreds to thousands of dollars, respectively, and do not include the additional course fees which reportedly cost upwards of A$2000 [20]. Two community members noted that despite experience in translating and interpretation, they do not have a license for translation because they cannot afford the fees [CL 04, 09]. “We just speak the language and we make sure that it’s right for the public” [CL 04]. While question remain on whether accreditation is necessary for all translated material, having someone else – especially someone from the affected community - to cast an eye over the translation to review it for accuracy *and* relevancy to the community was suggested. However, there are concerns that individuals voluntarily undertaking translations themselves could be misinterpreting data, misrepresenting information, or inadvertently changing meaning, signaling the engagement of NAATI certified translators is necessary to ensure the validity of translations. Finding the right balance of accuracy, speed, breadth of dialicts, and cultural knowledge appeared to be a priority for the community leaders – and that there needed to be a way forward to use both accredited and non-accredited translators during public health emergencies.

### 6. “Most of them are tuning into overseas stations”: Relying on overseas health messaging among CaLD communities in Australia

Reliance on accessing COVID-19-related health information from overseas was dominant theme among our community leaders. While some information of a general nature about COVID-19 – what is the virus, how does it spread, how to wash hands and socially distance - in community languages was useful; that heavy dependence on information from overseas overshadowed locally-relevant information about lockdown rules and government support, for example, leaving some CaLD community members in the dark about how COVID-19 was playing out in Australia. Representatives of Serbian, Middle Eastern and Indonesian communities, for example, expressed concern that elderly members of their communities were digesting COVID-19 information exclusively from overseas resources [CL 05, 11, 12, 15, 21]. Older generations of CaLD communities, in particular, may access their news from overseas newspapers and streaming TV services, missing critical local information. “There wasn’t proper communication, I have to say from an Australian perspective, what is happening here, what is available here? That’s the reality for those elderly [people]” [CL 11]. One community leader noted that as CaLD populations age, coupled with the heavier reliance on the internet and social media, some local community language newspapers have been declining in circulation [CL 12]. As a result, they attract less advertising dollars, and the commercial viability of these news outlets are compromised, cutting off an important source of local information for elderly migrant community members; hence the reach overseas.

Accessing overseas information it is not limited to just older people from CaLD backgrounds. Newly arrived migrants and refugees also needed better support to access information in Australia. “The government needed to collaborate a lot with community workers in order for this message to reach the community…When new migrants come here, they still watch the news in their own language…whatever they used to watch overseas”[CL 21; echoed by CL 03 as well]. Access to overseas media continues to occur amongst the general CaLD population as well. “Talking about the Arabic community, you’ll find that most of them are tuning into overseas stations. This is a problem we may have because the perception of COVID-19 is so different in Arab countries and we will never know the truth of the numbers…If we say one thing and then they’re watching another, the oversea channel and they get another message. It’s a conflicting situation for them” [CL 15]. In the context of a pandemic, bypassing local mainstream media in favour of overseas media meant critical public health information was not accessed by members of the CaLD communities, both young and old.

While some overseas information caused confusion amongst CaLD communities, it did have utility for some community organisations, especially when there was a paucity of translated materials in Australia. Community leaders acknowledged that they were actively seeking health information from overseas governments given material was already in the community language. “Because I speak Arabic, I have access to other Arabic countries’ media, and I found some material from the Saudi Arabia Health Department. I sent it to [a multicultural organisation] and to other people saying, ‘use it’ as a model of addressing the issue in Arabic. There is no need to reinvent the wheel if somebody did something good” [CL 19].

At the same time, some community leaders were cautious about accessing overseas information for fear it would not be relevant to the Australian context. “I access the translated version when I need to translate it, for example or to proofread it within my role…but normally the fastest thing is to access whatever you have. It’s always in English because the Arabic one could relate to the Middle East…I don’t want to give the wrong information while I’m reading something that is not related to Australia” [CL 04].

The concerns about CaLD community members accessing COVID-19 related health information overseas was not universally shared amongst our community leader interviewees. One participant acknowledged that while some people may not understand English, the vast majority were accessing information locally, especially through the daily State government press conferences during the height of the lockdowns. “Honestly, I don’t access any overseas information…I wouldn’t bother. What for? Why would they get information from overseas? Why can’t they get it here? Service New South Wales, if you can download their app for COVID, the information was there” [CL 07]. Awareness of the potential reliance on overseas information which may, or may not, be relevant to the local context needs to be a consideration of pandemic preparedness planning with CaLD communities going forward.

### 7. “You can’t reach them by the traditional way”: Harnessing social media

There was a resounding agreement amongst the community leaders about the growing importance of harnessing social media in disseminating critical public health messaging [CL 08, 09, 12, 15, 16, 20, 17, 21]. Mainstream media is no longer sufficient in reaching the majority of populations groups, especially CaLD communities. “You can’t reach them by the traditional way. If you want to reach them, you’re going to use platforms like Facebook, Instagram, and [messaging apps like] WhatsApp [or Signal or WeChat], whatever they use…If you want [to reach CaLD] communities, you’re going to have to use social media” [CL 21]. As noted above, traditional modes of community engagement, such as through local newspapers which may be produced in community languages, are slowly vanishing under the wave of social media which lowers circulation numbers leading to a disincentivisation of advertising revenue [CL 12]. During the pandemic lockdowns, community leaders had to shift face-to-face interactions to online platforms. Welfare and well-being checks would be conducted through text messaging, and translated recordings by community leaders and organisations about symptoms of COVID-19 and where to get tested, for example, would circulate on messaging platforms [CL 14]. “Everyone is so attached to their mobile phone for Facebook and WhatsApp. They can listen on WhatsApp and read through Facebook, and [other social media platforms] like that…The social media will be the best to reach them, from the young to the old” [CL 16, echoed by CL 08 and 21]. This community leader noted that one must not rely on social media in isolation, however, and that additional forms of communication, including flyers and radio, also have value in reaching those who do not access social media [CL 16].

Several community leaders used social media platforms and web-based messaging to receive information, filter it and forward the most important and relevant health information to various community groups [CL 08, 09, 10]. “I actually receive information from all sorts of different organisations, government and non-governmental organisations, and from the Senior Services Agency. I also share with them to my Facebook group so that people are on top of other things” [CL 09]. One community leader also noted that YouTube videos, in addition to other platforms, also contain important information in one’s own language and dialect and links can easily be forwarded community members and should be further levered by the government [CL 10]. “The Arabic translated messages that come from the government, from the Health Department, is not well-presented or well-distributed…I think [health messaging] should go to the Facebook…I think they should use the Facebook to send it to almost everybody or everywhere - something in Arabic so that people read it” [CL 12]. The importance of using social media to communicate information about the COVID-19 vaccine in particular was also raised by several community leaders. “Facebook is important, everybody sees it” [CL 12; echoed by CL 15, 16 and 21].

Community organisations are adapting to provide COVID-related information. One community group, in conjunction with a multicultural organisation in Western Sydney recorded several webinars in Arabic, such as “COVID-19 Updates and Hazards, COVID-19 Implications of Self-Isolation on Family Life, The Impact of Social Media Related to COVID, Improving Immunity and Treating COVID, and The Road to COVID-19 Vaccination” [CL 12], among others, which featured eminent doctors and health specialist to disseminate key COVID-19 information to the Arabic-speaking population of Western Sydney. These webinars were then recorded and posted on social media, such as YouTube and Facebook, and circulated, increasing their visibility across the community [CL 19].

While some community leaders expressed excitement at the possibilities social media brings in disseminating information, concerns were also raised by others related to the theme above – on ‘bridging the gap’ - about the volume of the messages circulating, and resources community organisations and individuals needed to sift through those messages to find the most relevant ones to convey to the broader community [CL 03]. Furthermore, while there are clear benefits in shifting additional resources into increasing the government’s social media engagement, there are trade-off with these platforms; in particular, the proliferation of false and misleading information. “People are relying on social media, which is selling all sorts of stories. We had to take it on into our own hands, actually do the work and talk the talk [to inform our community]. We did not see much from government [engagement with] the community” [CL 17].

There was some disagreement on whether the government’s outreach to CaLD communities was acceptable. One participant frankly noted that outreach, especially through social media, was absent. “I haven’t received one message about COVID-19 and vaccines [in my community language]” [CL 21]. Other participants, however, expressed satisfaction with the outreach and communication strategies employed by the government, remarking that the amount of pandemic-related information available across a number of platforms has been sufficient. “Everywhere you go, information is there…Published, posted, I’m seeing [information] everywhere on social media…I appreciate all these strategies.” [CL 18]. However, this participant did also caution against relying too heavily on print material. “If you want to hide any information from [certain] people, put it in writing, that is a saying” [CL 18]. Several other community leaders were critical of print documents as the dominant mode of disseminating information, noting bluntly that “people don’t read” [CL 3] and “no one reads that” [CL 11]. These community leaders hint that seeking alternative modes of delivery – such as through more targeted social media messaging - may be needed to reach those who avoid print material.

Where print and social media fail, the last resort is to engage one-on-one contact in-person or the telephone. “No one reads posters anymore and no one reads emails, the general emails. You have to either give me a call like this, a live person. I need to talk to a live person” [CL 11]. This is the most resource-intensive mode of communication given the costs associated with engaging a person to reach individuals, losing the broad reach of other messaging. “You don’t have the money for [hiring interpreters]. You have money for other stuff. It’s much cheaper to print the poster than to pay a person to go somewhere” [CL 11]. While it is clear that harnessing social media to disseminate critical public health information will need play a very significant part in future communication strategies, it will not be the *only* part of a strategy. The government will need to consider how to sunset some forms of communication that may not be relevant in a couple of generations as more and more people have significant internet use experience, but having a variety of communication platforms with both significant reach, and also one-to-one contact will still be required in the years to come.

### 8. “People really want to know the truth”: Challenges of communicating complex medical information to CaLD communities

The rapidly evolving public health information during the pandemic created unique sets of challenges for outreach efforts to the CaLD communities. Increasingly the challenge of providing clear, scientifically-based advice in relation to COVID-19 vaccinations emerged. The evolving data on the efficacy of the newly created COVID-19 vaccines, coupled with intense media scrutiny on the possible side effects and one’s own personal risk assessments, complicated the vaccine roll-out for the general population and CaLD communities alike. In discussing the challenges of communicating complex medication information to their community members, CaLD community leaders noted they initially struggled to access information about the vaccine that their community members wanted. While some of the technical information about the vaccine was available, concerned community members had additional questions about the safety and efficacy of vaccines – e.g. which vaccine is better? What are the side effects? [CL 08]. One community leader expressed disappointment in the public health communication, especially strategies to target multicultural communities. “I am a bit disappointed, because I consider myself to be relatively highly educated, and I had [feedback from people] who had problems with understanding what we’re up to [in the pandemic], or getting conflicting messages and thinking the government has been a bit slack in applying and implementing policies, and then changing their mind. If I’m finding that hard, and I do feel for the rest of the community, or for people who don’t have that certain level of exposure to education” [CL 15, echoed by CL 17]. While broad understanding of the evolving nature of the pandemic was a challenge for most of the population, information about vaccines represented a flashpoint for misinformation.

One community leader emphasised that people want to be informed about the vaccine. “The people really want to know the truth” [CL 20]. While noting the government was initially not forthcoming with information about the vaccine targeting CaLD populations, community organisations “tried to explain to our community the real fact of having vaccine” [CL 20]. However, feeling ill-equipped to answer some basic questions about health messaging, community members turned away from community organisations and towards other sources of information on the internet, unhinged from data or facts. The resulting conclusion led to community leaders fielding a significant amount of phone calls from concerned community members seeking more information (see CL 20 in the fourth theme above). Several community leaders have grappled with how to reduce the spread of false and misleading information about COVID-19 generally, and the vaccine, specifically. Some community leaders noted that CaLD community members – older generations in particular - carry skepticism of government information, a by-product of some migrant and refugee backgrounds previously living under authoritarian rule where governments could not be trusted. However, while most young people in the community (20 years and younger) know English and can access information easily, older, first-generation migrants and refugees, especially those born under dictatorships, “don’t believe what the government says…Everybody’s suspicious of everything” [CL 12]. Such distrust in government adds another layer of complexity in reaching older people from CaLD backgrounds, on top of several challenges already highlighted above.

Beyond providing basic health information about vaccines, community leaders suggested that the COVID-19 vaccine roll-out has not been fair and equitable, and CaLD communities did not receive enough support in accessing the vaccine [CL 15, 20]. “The leaders have been very slow to accept when we have to encourage people to do the right thing and get vaccinated. This my impression, and now we are trying to remedy the situation” [CL 15]. This community leader went on to note that the government’s COVID-19 response was inequitable. “We can see the discrimination, we can see the contradiction, the conflict in the messages we have been receiving. Let alone assistance to get the vaccines. No, it hasn’t been helpful as such” [CL 15].

This community leader suggested that approaching General Practitioners from multicultural communities should have been a priority in the vaccine roll-out. “The GPs, we should have perhaps started with some multicultural, ethnic background GPs to get them on board, to say to people they can go to them [for vaccine information]. It has been very, very vague messaging. No, I think this is one thing that we missed” [CL 15]. Community leaders suggested CaLD doctors could strike the right balance of explaining public health information scienfically, but not too technically, while providing clear, simple advice [CL 15, 22] [21].

When it comes to COVID-19 vaccine information, community leaders had one key message: that vaccine information should come from well-known bilingual medical or health professionals from the community, who have the relevant public health expertise, and are credible sources of information [CL 12, 14, 22]. “I believe that community leaders should not go and just pass on messages related to health…the only people that [pass on public health messages] efficiently are the community doctors themselves” [CL 22]. While other community members, especially respected elders and faith-based leaders, can support these efforts, their core business is not health-related messaging and as such, should be secondary – albeit important - sources of information for the community given their elevated status, after health professionals lead the advocacy efforts. “[A religious leader] does not have health expertise whereas the message is more credible if it comes from a health professional” [CL 14]. There were also suggestions that faith-based leaders may have concerns about the vaccine, and thus are not best-suited to act as health promoters. “[N]one of [the religious leaders] have been vaccinated…If they don’t believe [in the vaccine] themselves, how can they spread the message? [CL 14]. Such comments call into question the possible role that community leaders and faith-based leaders could play as spokespersons in vaccine information dissemination strategies, which governments have proposed, and in the case of Victoria, implemented through a Vaccine Ambassadors program [22]. “I still believe health should deliver that community message through us like bilingual health workers to deliver the messaging in their first language in the community” [CL 14].

At the same time, health professionals are not universally supportive of the COVID-19 vaccine, with some expressing skepticism about the benefits of the vaccine, and focusing on the potential harm. It was reported in some communities that some bilingual doctors add fuel to the skepticism by openly suggesting to patients they “don’t know what’s in the vaccine and the drawbacks” [CL 12]. While such anecdotes deserve greater scrutiny, it does flag that governments and community organisations must be aware of skeptics within the medical/health profession, as this could diminish community reach about the scientific basis and efficacy of the vaccine.

Some community members offered solutions to addressing the communication challenge in disseminating vaccine information to CaLD communities. In relation to addressing vaccine hesitancy among CaLD community members, one community leader noted that providing space for one-on-one consultations with a health practitioner, accompanied by an interpreter, could have helped to address their concerns about the vaccine [CL 11]. Another community member suggested simplifying the messaging. “Not many people really understand what is this vaccine? But by explaining in a simple way and in simple language, they could understand it very easily” [CL 16]. Another community member noted at the time of interviews as the vaccine roll-out for the Delta variant (Aug 2021) that the government was ramping up its public health engagement with CaLD communities. “I didn’t get anything yet about the vaccine…[but] they started saying that we’ll be expecting lots of work because we are the clinical worker with languages skills, I think 110 languages. We’ll be expecting information about the vaccination…That’s why I’m dealing directly with doctors when I need information.” [CL 04]. It was clear from the community leaders that clear, scientifically-sound public health information delivered by trusted health practitioners representing CaLD communities was the most effective mode of outreach.

## DISCUSSION

The overwhelming message from community leaders was a sense of shared responsibility between their organisations and the government in communicating pertinent and accurate COVID-19 related information to CaLD communities. Community leaders expressed a sense of duty to keep their community members safe. While acknowledging this shared responsibility, however, there were significant costs shouldered by community leaders, other individuals and organisations related to resources and time that need to be acknowledge by government going forward in preparing for future disease outbreaks. Further public health information campaigns that target CaLD communities have several considerations for improving their translating and interpreting capacity. There are several ideas emerging from discussion with community and faith-based leaders – the information intermediaries - that governments should consider: 1) better communication between governements and CaLD community organisations; 2) the need for agility; 3) a suite of modes of communication; 4) reinvesting in local public health units; 5) a health ambassadors’ model; 6) a hybrid model of translators/interpreters; and 7) reimagining vaccine information campaigns.

### Better communication between governments and CaLD community organisations

A tension emerged from our discussion with community leaders, suggesting the government was placing significant responsibility on community leaders/organisations to get the public health messaging to communities. To address the discord between government and the community, one leader suggested more communication between government representatives and community leaders is required. “When someone [well known] from the community and from the government sit down together, they can talk. I believe this is how they can deliver [public health messaging] easier. This is what we’re trying to do…we complete, a little bit, the gap, in communicating real information with our community” [CL 20]. A more collaborative approach between government and CaLD communities was echoed by other leaders. “I think it’s a joint responsibility. The Health Department with all the experts they have to provide the information and rationalise, and then the main media outlet, like newspaper and the television and other things, as well as the media targeting the multicultural community. I’m sure also that the ethnic organisations or the multicultural organisation have a bigger role to play once they are supported and given the right information and given the right tools to spread the information” [CL 19]. The government expected community leaders to disseminate key public health messages, and the vast majority were willing partners in the efforts to inform their people; however, while there was a shared responsibility, there was not a always a shared cost. While community leaders were prepared to ‘bridge the gap’, appropriate funding and support for costs incurred for the significant time and resources required was an issue repeatedly flagged by the participants.

### Need for agility

Agility is not a term typically used to describe government action. However, based on the general lesson learned about the pandemic [23], and insights from the community leaders throughout this paper, it is clear that pandemics of the nature and scale of COVID-19, the ability of government to rapidly shift and pivot based on current data is required for an effective public health response. Importantly, however, in doing so, is having effective mechanisms in place to bring the community along with it, especially those from CaLD communities where English proficiency is limited and trust in governments is strained.

It was clear in the early stages of the pandemic that the public was not prepared for the evolving nature pf public health advice (e.g. not wearing masks at the beginning of the pandemic to safeguard stockpiles for health care workers and patients, to recommending mask wearing by all members of the public when outside one’s home). In future pandemics, governments need to make clear with the public at the outset that health advice is not static and will evolve and change as new data emerges and resources change. In doing so, the community can be prepared for shifting advice day to day when necessary. Alerting CaLD communities, especially those with low English language proficiency and less community infrastructure, and the community organisations that serve them, about the possibility of evolving public health advice will be an important feature in future pandemic preparedness.

### Suite of modes of communication

One of the strong messages from the community leaders is that a suite of communication strategies is needed to reflect the diversity of CaLD communities, the generational differences in accessing internet-related information, and those with low levels of English proficiency. While public health messaging through TV, radio, newspaper and posters must remain part of the modes of communication strategies, it is clear from our community leaders that CaLD communities are increasingly engaging with a wider variety of social media platforms to access and share pandemic-related information. Social media platforms such as Facebook, YouTube, and WhatsApp were widely cited amongst our participants as popular modes of communication. The rapid availability of public health information through these platforms was critical to informing the CaLD communities about the pandemic. However, there were several pitfalls cited by the community leaders in using social media. First, the source of content was an important consideration. As noted in the theme above ‘bridging the gap’, community members readily stepped in to translate and interpret critical public health information and post these resources on various social media sites. The organisations often did so voluntarily, though this additional engagement strained many organisation’s resources. Second, access to social media meant the general public, including CaLD communities, had easy access to a mirage of false and misleading COVID-19 information. The susceptibility to false information led to further strains within community organisations to field additional questions about the pandemic. It also lead to concerns about vaccine hesitancy and created challenges to getting the general public on board for mass COVID-19 vaccine campaigns. Finally, another challenge with social media was the ease of accessibility to overseas pandemic information that may not have been applicable to the Australian context. While some CaLD communities may have tuned into overseas TV and radio programs, the rapid exchange of overseas information in their community language coloured their perception of the pandemic locally. Reviewing and revising public health messaging strategies will need to place greater attention on the strength of engaging with social media, while also acknowledging the pitfalls of the platform and mitigating against negative effects [24]. At the same time, as generations evolve, there may need to be a sunsetting of some more traditional modes of communication – translated newspapers and radio programs, for example – in favour of internet-based communication strategies.

### Reinvesting in local public health units

One of the important early lessons from the pandemic in Australia was that early and sustain investment in local public health units made a material difference in how the pandemic play out in real time, particularly when comparing COVID-19 rates in NSW and Victoria [25,26]. Prior to the pandemic, NSW’s public health approach was more decentralised, where local public health units had significant command of their populations. By contrast, the Victorian public health approach was centralised, leaving public servants based in Melbourne to make decisions across the state, where knowledge of the local population and area was limited. This dichotomy in public health approaches across State/Territory lines was a critical difference in the trajectory of the pandemic [27]. Greater research on how the various local public health responses to CaLD communities specficially across State/Territory lines is warranted.

The CaLD community leaders emphasised the need for governments to reinvest in community engagement, in knowing one’s population [28]. This could take a variety of forms, including increased investment in local public health unit outreach with CaLD communities, formalising more partnerships with community organisations, fostering regular engagement with community organisations both during the pandemic and non-pandemic periods, and ensuring a very clear understanding of the cultural and linguistic diversity *within* the community so that no one gets left behind in public health emergency communication strategies. Such investments should not rest solely on the shoulders of public health, but have wider appeal to other emergency situations, such as fires and floods, where rapid transmission of urgent information is simply and clearly communicated to the affected communities, in a language they understand.

### Consider a health ambassadors model

The community leaders clearly suggested that public health information should come from health practitioners, who have the medical and/or public health knowledge, high levels of cultural sensitivity, and the trust and respect of the local community. Some community leaders expressly stated that investment in one-on-one engagement is preferable, even if not financially practical. While resource intensive, governments could consider a variation of a model used in Victoria to support vaccine uptake, called Vaccine Ambassadors [22]. These ambassadors were trained to travel door to door to engage with local community members and have conversations about the vaccine, answering questions about safety and efficacy, and where to access vaccination clinics. Further research on the impact of these vaccine ambassadors and the potential to broaden their scope to ‘health ambassadors’ could be a way forward to bridging government public health with CaLD communities.

### Hybrid model of translators/interpreters

The volume and urgency of public health information disseminated to the public led to community leaders and organisations to step in to fill the gap in reaching CaLD communities. In doing so, many community leaders noted that they did so given they had the connections to the community and the language ability to translate and interpret the public health messaging. However, some acknowledged that they did not have the official NAATI accreditation which recognises trained translators and interpreters. A tension emerged between the need to have accurate translations and the urgency in which information need to be disseminated. There were concerns that individuals voluntarily undertaking translations themselves could misinterpret data, misrepresenting information, or inadvertently changing meaning, signaling the engagement of NAATI certified translators is necessary to ensure the accuracy of translation. While it is imperative to safeguard that accreditation, there may need to be a hybrid model where governments can call on NAATI accredited translators and interpreters, but also informal translators networks, to disseminate critical health information.

### Reimagining vaccine information campaigns

One of the most important lessons to be learned from this COVID-19 pandemic is how to more effectively develop and implement vaccine information campaigns to reach CaLD communities. Understanding the effectiveness of public outreach to inform community members and support their decision-making process is critical to future public health emergencies. Community leaders emphasised the need for increased outreach to multicultural communities to support their decision-making about the vaccine [CL 16]. While there may not be one single approach that will best support CaLD communities, offering them factual, scientifically-based information that is understandable to the layperson is an important component of any strategy. “The only thing we have to do is [explain] how to increase our immune system in our body to make ourselves more healthy” [CL 16]. As noted above in the other themes, harnessing social media, printing flyers, promoting multicultural health communication on radio and TV, all need to be part of a multi-prong strategy of vaccine information outreach.

To support adoption of government measures including a future COVID-19 vaccine, it is essential that people can access to, understand, and trust the messaging about COVID-19 vaccines, that materials meet the health literacy needs, and communication approaches are culturally contextualised. Given the ongoing nature of this pandemic, and the possibility of regular updates to the COVID-19 vaccine going forward, there is an urgent need to strengthen communication efforts with CALD communities to ensure that community engagement is enhanced and there is appropriate ongoing adherence with current response strategies and the introduction of a future COVID-19 vaccine.

## Conclusion

Culturally and linguistically diverse communities are an important part of the Australian population and it is imperative that their needs are met as part of the Australian response to ongoing and future pandemic responses. Unfortunately, evidence suggests that CaLD communities bore a significant burden in the Australian response to the COVID-19 pandemic [29]. It is imperative of government, academia, health bodies, and community and faith-based organisations to heed the lessons learned and ensure that CaLD communities do not get left behind in future pandemic and public health emergencies, and communication strategies that include a diverse and flexible approach to reaching CaLD community members should consider new modes of engagement beyond translation and interpretation services.

## Data Availability

The datasets generated during and/or analysed during the current study are not publicly available due to the sensitive nature of the topic raised during the interviews, but are available from the corresponding author upon reasonable request.

## Declarations

### Ethics approval and consent to participate

The Human Research Ethics Advisory Panel at the University of New South Wales reviewed and approved this study (HC200776). All of the methods used in this study were performed in accordance with the relevant guidelines, outlined by The National Statement on Ethical Conduct in Human Research, published by the Australian Government. Informed verbal consent was collected from all participants.

### Authors’ contributions

HS was responsible for the design of the study, for conducting data collection and analysis; BHR, AH, IA, AM and LW supported the development of the study and interpretation of the findings. EW supported the analysis of the transcripts.

## Acknowledgements

The research team wish to thank the participants for their support of this study.

## Appendix 1: CORE-Q Consolidated Criteria for Reporting Qualitative Research

| No | Item | Description |
| --- | --- | --- |
| **Domain 1: Research Team and Reflexivity** |  |  |
| **Personal Characteristics** |  |  |
| 1 | Interviewers | HS, BHR |
| 2 | Credentials | The two interviewers between them hold Bachelor, Master and PhD qualifications in behavioural science, social work, social research, medicine, and public health. |
| 3 | Occupation | Academic staff (HS, BHR, AH, IA, AM, LW, EW) and public health practitioner (LW) |
| 4 | Gender identity | Female (HS, AH, AM, LW, EW), male (BHR, IA). Two participants identify as from CaLD backgrounds (IA, AM) |
| 5 | Experience and Training | Experienced researchers and interviewers, have undertaken several qualitative studies, trained in interviewing and qualitative analysis. |
| **Relationship with participants** |  |  |
| 6 | Relationship established | None of the interviewees were known to the interviewers prior to the interviews. |
| 7 | Participant knowledge of the interviewer | The participants in this study were representatives of the various community and faith-based organisations from across Australia. The interviewers were not known to participants before the interviews were conducted. |
| 8 | Interviewer characteristics | Established researchers with experience in vaccination, public health, health services research, primary health care, and culturally diverse health research. Collectively the interviewers have more than 40 years of research experience. |
| **Domain 2: Study Design** |  |  |
| **Theoretical Framework** |  |  |
| 9 | Methodological orientation and theory | Exploratory |
| **Participant Selection** |  |  |
| 10 | Sampling | Purposive and snowballing, with participants sought across Australia. Participants were also asked to identify other participants to be interviewed. |
| 11 | Method of approach | Emails which included information on the study, participant information and consent forms. |
| 12 | Sample size | 30-40 |
| 13 | Non-participation | 11 participants didn't respond. |
| **Setting** |  |  |
| 14 | Setting of data collection | Conducted via online videoconference platform and telephone |
| 15 | Presence of non-participants | No non-participants were present for the interviews |
| 16 | Description of sample | Participants were representatives from CaLD community organisations and groups from New South Wales, Australia. |
| **Data Collection** |  |  |
| 17 | Interview guide | Provided in advance upon request |
| 18 | Repeat interviews | No |
| 19 | Audio/visual recording | Audio recording |
| 20 | Field notes | Limited reflective notes were captured following interviews. These notes were not included in the qualitative analysis. |
| 21 | Duration | Ranged between 20 minutes to 60 minutes |
| 22 | Data saturation | Purposeful sampling to ensure range of experiences from across different cultural groups were captured and described. As such sampling continued past basic conceptual/thematic saturation to ensure a range of cultural perspectives were included. |
| 23 | Transcripts returned | No |
| **Domain 3: Analysis and Findings** |  |  |
| **Data Analysis** |  |  |
| 24 | Number of data coders | Initial coding was done by HS and EW. BHR contributed to development and refining concepts identified. Codes, concepts and higher order themes were discussed by all authors. |
| 25 | Description of the coding tree | No, not necessary given the focus of the analysis is largely descriptive rather than being theory-driven. |
| 26 | Derivation of themes | Codes and concepts were derived through open coding of interview transcripts, these were then tested and consolidated into higher order themes. |
| 27 | Software | NVivo |
| 28 | Participant checking | No |
| **Reporting** |  |  |
| 29 | Quotations presented | Yes, selectively to illustrate findings |
| 30 | Data and findings consistent | Yes |
| 31 | Clarity of major themes | Yes |
| 32 | Clarity of minor themes | No, the focus in this paper is on major analytic findings and higher-order themes, given the focus of the study. |

